# Improved Alzheimer’s disease versus frontotemporal lobar degeneration differential diagnosis combining EEG and neurochemical biomarkers

**DOI:** 10.1101/19009316

**Authors:** Jorne Laton, Jeroen Van Schependom, Joery Goossens, Wietse Wiels, Anne Sieben, Peter Paul De Deyn, Johan Goeman, Johannes Streffer, Julie van der Zee, Jean-Jacques Martin, Christine Van Broeckhoven, Maarten De Vos, Maria Bjerke, Guy Nagels, Sebastiaan Engelborghs

## Abstract

**Introduction:** Distinguishing between two of the most common causes of dementia, Alzheimer disease (AD) and frontotemporal lobar degeneration (FTLD), on clinical diagnostic criteria alone has poor diagnostic accuracy. Promising tools to increase the diagnostic accuracy of AD are the use of cerebrospinal fluid (CSF) biomarkers and electroencephalography (EEG), which is being investigated as a diagnostic tool for neurodegenerative brain disorders. In this study, we investigated the utility of EEG-based biomarkers in comparison and in addition to established neurochemical biomarkers in the AD versus FTLD differential diagnosis.

**Methods:** Our study cohort comprised 37 AD and 32 FTLD patients, of which 19 AD and 11 FTLD had definite diagnoses. All these participants had CSF biomarker analyses resulting in four neurochemical (NCM) biomarkers (Aβ_1-42_, T-tau, P-tau_181_ and Nf-L) and underwent 19-channel resting-state EEG. From the EEG spectra, dominant frequency peaks (DFP) were extracted in four regions resulting in four dominant frequencies (in left-temporal, frontal, right-temporal and parieto-occipital regions). This yielded a total of eight features (4 NCM + 4 EEG), which we used to train and test a classifier and assess the diagnostic value of the markers separately (using only the NCM or EEG subset) and combined.

**Results:** The classification accuracies were much higher when training and testing on the definite subgroup than on the whole group. Furthermore, we found that the NCM feature subset allowed a better accuracy than the EEG feature subset, both when training and testing on the whole group (NCM 82% vs EEG 72%), as on the definite group only (92% vs 86%). Using both feature subsets together increased the accuracy to 86% in the whole group and 94% in the definite subgroups. Another interesting finding was the presence of two spectral peaks in a considerable number of patients in both groups.

**Conclusion:** Combining EEG with neurochemical biomarkers resulted in differential diagnostic accuracies of 86% in clinically diagnosed AD and FTD patients and of 94% in patients having a definite diagnosis. Furthermore, we found evidence that the slowing down of the dominant EEG rhythm might be a gradual appearance of a slow rhythm and fading out of the normal ground rhythm, rather than a gradual slowing down of the ground rhythm. Finally, we have discovered two differences in the occurrence of the dominant EEG frequency: people lacking a clear dominant peak almost all had definite AD, while people with two peaks more often had FTLD. These unexpected but interesting findings need to be explored further.

## 1. Introduction

Distinguishing between two of the most common causes of dementia – namely Alzheimer disease (AD) and frontotemporal lobar degeneration (FTLD) – on clinical diagnostic criteria alone has poor diagnostic accuracy, with high numbers of autopsy-confirmed cases contradicting the clinical diagnosis (Elahi and Miller, 2017; Engelborghs et al., 2008), mainly due to overlap in clinical presentation, especially in the early disease stages.

A promising tool to increase the diagnostic accuracy of AD is the use of cerebrospinal fluid (CSF) biomarkers (Niemantsverdriet et al., 2017). During the past decades, a lot of progress has been made to improve AD diagnosis, resulting in biomarker-based research diagnostic criteria, also for its prodromal stage of mild cognitive impairment (MCI) due to AD. Therefore, revised criteria of both the National Institute on Aging/Alzheimer’s Association (NIA-AA) and the International Working Group (IWG) include AD CSF biomarkers in the clinical diagnostic work-up (Albert et al., 2011; Dubois et al., 2014; McKhann et al., 2011).

Analysis of the core AD CSF biomarkers (β-amyloid peptide composed of 42 amino acids (Aβ_1-42_), the Aβ_1-42_ /Aβ_1-40_ ratio, total tau protein (T-tau) and phosphorylated tau at threonine 181 (P-tau_181_)) can help to differentiate between AD and non-AD dementias like FTLD, but they cannot be used to confirm a non-AD dementia (Niemantsverdriet et al., 2017). Moreover, pathological values of these core AD CSF biomarkers can be seen in non-AD dementias, which might lead to possible misinterpretation of the biomarker results in absence of clinical information. Indeed, both Aβ_1-42_ and T-tau can be detected at intermediate levels (in between normal control and abnormal AD values) in non-AD patients, also in FTLD. CSF P-tau_181_ is a more specific marker for AD and is of help for AD versus non-AD dementia differential diagnosis (Struyfs et al., 2015). However, in order to improve the discriminatory power for the differential diagnosis of dementia, additional markers, more specific to the non-AD dementia are valuable. As CSF levels of neurofilament light chain (Nf-L) were significantly higher in FTLD compared to AD and controls, there is an added value for Nf-L in the differential diagnosis of FTLD (Goossens et al., 2018).

Electroencephalography (EEG) is an easily accessible, non-invasive, inexpensive technique, and is already being investigated as an adjunctive investigation in dementia (Ferreira et al., 2016). We have previously shown that EEG maxpeak frequency is an easy and useful measure with an added value in the differentiation between AD and FTLD, reaching a diagnostic accuracy of 78.4% (Goossens et al., 2017).

In this study, we investigated the utility of EEG-based biomarkers in comparison and in addition to established neurochemical biomarkers in the AD versus FTLD differential diagnosis.

## 2. Methods

### 2.1 Study participants

The study cohort comprised 37 subjects with probable (n=18) or definite (n=19) dementia due to AD, and 32 probable (n=21) or definite (n=11) FTLD patients (Table 1). Patients were selected from the Memory Clinic of Hospital Network Antwerp (Engelborghs et al., 2006, 2003). To ensure a high certainty level of dementia subtypes for patients without definite diagnosis, only patients with extensive clinical follow-up were included. All patients underwent (among others) neuropsychological testing including Mini-Mental State Examination (MMSE), and had core AD CSF biomarker analyses (Aβ_1-42_, T-tau, P-tau_181_) in cerebrospinal fluid (Somers et al., 2016). Clinical diagnosis of probable AD was based on NIA-AA (McKhann et al., 2011) and IWG-2 criteria (Dubois et al., 2014) and included these biomarkers. Diagnosis of probable FTLD was based on criteria described by Neary (Neary et al., 1998). Subgroups of definite dementia patients were defined by genetic carrier status and/or post-mortem confirmation of brain pathology (Mackenzie et al., 2011; MacKenzie et al., 2010; Montine et al., 2012). This study was approved by the ethics committee of University of Antwerp (Antwerp, Belgium).

**Table 1:**
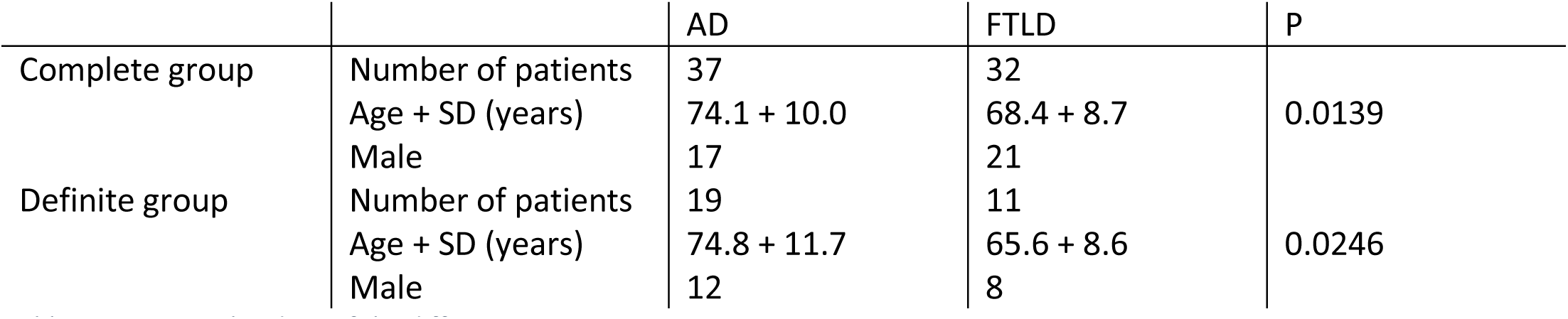
Demographic data of the different patient groups.

### 2.2 Neurochemical (NCM) biomarkers

Lumbar puncture, and CSF and blood sampling and handling was performed according to a standardized protocol (del Campo et al., 2012; Engelborghs et al., 2017). All CSF and blood samples were stored at the IBB Biobank in polypropylene vials at 80°C until analysis.

CSF biomarker levels were quantified using commercially available single-analyte ELISA kits (one kit lot each), strictly following the manufacturer’s instructions (INNOTEST β-Amyloid(1-42), INNOTEST hTau-Ag and INNOTEST Phospho-Tau(181P) from Fujirebio Europe, Belgium; and Nf-L from UmanDiagnostics. All samples were run in duplicate, blinded for diagnosis. Intra-assay coefficient of variation was below 10% for all analytes.

### 2.3 Electroencephalographic (EEG) markers

#### EEG recordings

EEG data were recorded using OSG digital equipment (BrainLab/BrainRT) with the international 10–20 system used for electrode placement. ECG was recorded in a separate channel. Recordings were exported in EEGLab format (Delorme and Makeig, 2004) for offline analysis and each file contained continuous data in 19 channels. During recording, subjects were seated upright and were asked to alternate between eyes closed and eyes open to stay awake. EEG data was processed manually using BrainRT. Artefact-free EEG during the eyes-closed condition was flagged. This flag consisted of start latency of the useable part and its duration, both in milliseconds. No epileptiform activity was observed in any of the EEG recordings. The EEGs were re-referenced to average.

#### Epoch extraction

In our previous study on this dataset (Goossens et al., 2017), we extracted two-second epochs. This proved to be too coarse to detect smaller frequency differences. Therefore, we doubled the epoch length to four-second epochs from each EEG and selected six epochs (the minimum over all patients) equally spread over all available epochs of that patient. This epoch length ensured retaining most of the artefact-free signal, while also allowing a spectral resolution of 0.25 Hz. We did not increase the epoch length further, as this started to increase loss of artefact-free EEG. With longer epochs, chances increase that certain continuous parts of clean EEG are not long enough to contain a number of epochs that accounts for the same total time.

#### Transformation to frequency spectrum

We used Welch’s power spectral density estimate (Welch, 1967). In Matlab (MATLAB, 2019), this method is implemented with integrated support for windowing and based on the built-in fft (Fast Fourier Transform). Instead of computing the spectrum on each epoch separately, we concatenated the epochs back into a ‘continuous’ signal. We aligned the windowing exactly with the epoch boundaries, by using a Hamming window with a length equal to the epoch length and the overlap between windows equal to zero. This resulted in a smooth, average spectrum with much smaller roll-off than a rectangular window.

To increase the resolution, we padded the epochs with zeroes, by setting the nfft parameter of Matlab’s built-in pwelch function to a multiple of the epoch length, thus increasing the resolution by the same factor. Padding makes the resolution higher, but does not add any more information. The extra points in the spectrum are estimated by sinc interpolation between the original points. For our purpose – detecting peaks in the spectra – this was perfectly adequate.

#### Dominant frequency peak extraction

Dominant frequency peaks (DFPs) were detected within the interval [5-15] Hz in every channel. A peak was defined as a point of which both neighbouring points have lower amplitudes. In case multiple peaks were detected, only the peak with the highest amplitude was retained, and this in each channel.

To reduce redundancy, balance the number of neurochemical and EEG features and deal with the fact that some channels did not always exhibit peaks, we summarised the peaks found over all channels into four regions. To do this, we fine-tuned the algorithm from our previous study (Goossens et al., 2017) to detect the DFP with the highest amplitude over the channels within a specific region, as defined in table 2. This procedure was also helpful in avoiding missing data, as only one channel per region needed to show a clearly distinguishable peak. The resulting dataset contained the frequency of every region’s DFP, for each subject.

**Table 2:** Region definitions.

| Region label | Electrodes |
| --- | --- |
| F (frontal) | Fp1, Fp2, F3, Fz, F4 |
| P (parietal and occipital) | P3, Pz, P4, O1, O2 |
| L (left temporal and central) | F7, T3, C3, T5 |
| R (right temporal and central) | F8, T4, C4, T6 |

### 2.4 Classification

#### Feature sets

We used three feature sets: one subset containing the four NCM markers, one subset containing the four EEG markers and one set of eight features containing both the NCM and EEG markers. Each of these sets was used for training and testing the classifier algorithm, such that we could compare the performance of the subsets and verify if combining them improves the accuracy.

#### Cross-validation

Training and testing were done using cross-validation. Features (NCM and/or EEG markers) were collected into one dataset, which was then randomly divided into ten subsets or folds with the same proportion of the two groups (AD and FTLD) as in the complete dataset. We applied tenfold cross-validation: nine folds were used for training the classifier, while the remaining fold was used to test the trained classifier. This step was repeated until each fold had been used exactly once for testing. The test results for each fold were then combined into a total estimate of the classifier’s performance.

We repeated tenfold cross-validation 100 times, which allowed for the calculation of the mean and standard deviation of the classifier’s performance indices: sensitivity, specificity, positive predictive value (PPV), negative predictive value (NPV) and percentage correctly classified (PCC, also called accuracy). The sensitivity and specificity respectively show the ratio of correct positives over actual positives and correct negatives over actual negatives. The positive and negative predictive values respectively show the ratio of correct positives over all instances classified as positive and negatives over all instances classified as negative.

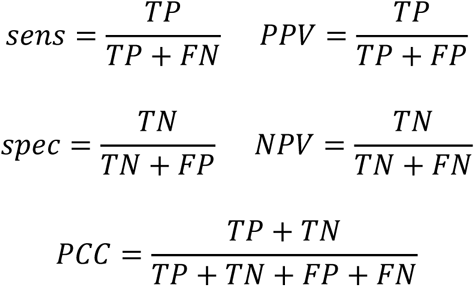

With TP = True Positives, correctly classified positives, TN = True Negatives, correctly classified negatives, FP = False Positives, negatives incorrectly classified as positives, and FN = False Negatives, positives incorrectly classified as negatives.

#### Classifier algorithm: Random Forest

A Random Forest is a collection of randomly generated decision trees. For each of these individual trees, a subset of features is selected at random from the complete feature set and then the tree is built. This resolves a major problem of decision trees: training a decision tree is difficult when there is a large number of features. After a fixed number of trees has been built, the prediction is based on majority voting of all trees. Most of these trees will be useless and giving random responses, but on average these will cancel each other out, which results in only the relevant trees adding to the prediction. We used the function randomForest of the R package “randomForest” (Breiman, 2001) with number of trees set to 500 and default parameters for this classifier.

## 3. Results

### 3.1 Age as potential confounding factor

Since our groups and subgroups significantly differed in terms of age, we calculated the correlation between age and T-tau, as well as between age and Nf-L.

In most studies, these neurochemical parameters are found to be correlated, but table 3 shows that this was not the case in our dataset, which might suggest that our study was underpowered to detect this correlation.

**Table 3:** Correlations between Age and Tau, and Age and NfL.

| Age with | Correlation | Significance |
| --- | --- | --- |
| Tau | 0.0294 | 0.8102 |
| Nf-L | 0.0715 | 0.5591 |

### 3.2 Data check – special spectra

When we verified the validity of our spectral peak detection algorithm, we found three spectrum categories and listed them in table 4. We also noticed that AD spectra were generally noisier and had lower amplitude than FTLD spectra. In a small number of patients, mainly AD, only in a few channels a dominant peak was found. However, these peaks were often not very pronounced in these patients: less than double the spectral average.

**Table 4:** Number of spectral peaks.

| Group | Weak peak and only in a few channels | 1 clear peak in most channels | 1 clear peak and 1 extra peak in several channels |
| --- | --- | --- | --- |
| AD | 7 | 24 | 6 |
| AD – definite | 6 | 12 | 1 |
| FTLD | 2 | 20 | 10 |
| FTLD – definite | 0 | 6 | 5 |

In a considerable number of participants, mainly FTLD, we detected two dominant EEG peaks in the spectrum at around 2Hz from each other. Of these two, the peak with the highest frequency was usually in the (healthy) alpha range, around 9Hz, and the other was usually around 7Hz. The two clearest examples of spectra with two peaks in either group (AD patient on the left, FTLD right) are shown in figure 1. As a side note, they both had a non-definite diagnosis.

**Figure 1:**
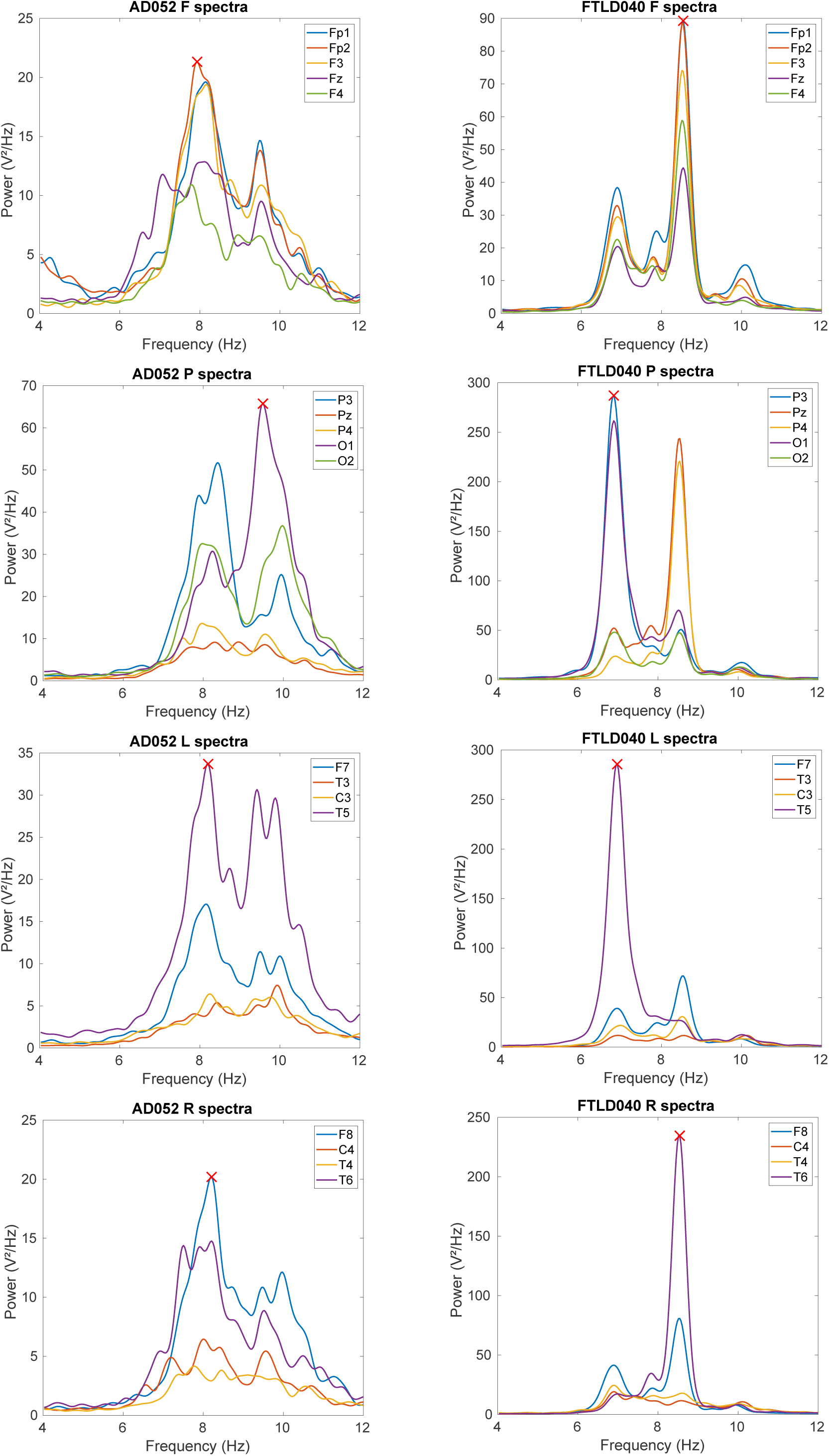
Two examples of patients who have two peaks in their spectrum. In each region, the algorithm is able to extract the best dominant peak candidate, which is marked with a red cross. Left column: AD patient; right column: FTLD patient. Top row: frontal (F); second row: parietal-occipital (P); third row: left-temporal (L); bottom row: right-temporal (R).

Looking at definite diagnosis only, no FTLD patient with a weak peak spectrum remained, while almost none of the AD patients with a double peak remained. Eventually, we found that our spectral peak detection algorithm was also robust against these special cases of only a few peaks, or two peaks in the spectrum, and always extracted the most logical one.

### 3.3 Classification

Tables 5 and 6 show classification results respectively in the complete and the definite group. Table 5 shows that the diagnostic accuracy of the NCM biomarkers result in a good accuracy. The EEG features are behind, but reasonable. Cross-validating on NCM and EEG features together yielded a better accuracy than on NCM or EEG individually.

**Table 5:** Classification accuracies cross-validating on the complete group.

| Features |  | Acc | Sens | Spec | PPV | NPV |
| --- | --- | --- | --- | --- | --- | --- |
| NCM | AV | 0.816 | 0.840 | 0.788 | 0.821 | 0.810 |
|  | SD | 0.013 | 0.012 | 0.020 | 0.015 | 0.013 |
| EEG | AV | 0.722 | 0.734 | 0.709 | 0.745 | 0.698 |
|  | SD | 0.023 | 0.031 | 0.035 | 0.024 | 0.027 |
| NCM & EEG | AV | 0.856 | 0.910 | 0.793 | 0.836 | 0.885 |
|  | SD | 0.021 | 0.028 | 0.026 | 0.019 | 0.032 |

**Table 6:** Classification accuracies cross-validating on the definite group only.

| Features |  | Acc | Sens | Spec | PPV | NPV |
| --- | --- | --- | --- | --- | --- | --- |
| NCM | AV | 0.917 | 0.937 | 0.882 | 0.934 | 0.894 |
|  | SD | 0.039 | 0.042 | 0.086 | 0.041 | 0.065 |
| EEG | AV | 0.863 | 0.905 | 0.791 | 0.882 | 0.830 |
|  | SD | 0.033 | 0.033 | 0.044 | 0.024 | 0.055 |
| NCM & EEG | AV | 0.937 | 0.979 | 0.864 | 0.926 | 0.962 |
|  | SD | 0.025 | 0.027 | 0.048 | 0.024 | 0.049 |

Including only the participants with a definite diagnosis greatly improved the classification accuracy. Table 6 reveals the same trend in which the NCM features already perform well, but the accuracy increases upon adding the EEG parameters, which are slightly behind on their own. We observed the same effect of increased accuracy in the definite group in our previous study focussing on EEG (Goossens et al., 2017).

## 4. Discussion

We wanted to investigate the value of neurochemical biomarkers and EEG-based markers to distinguish between the two most common causes of dementia that can still not be diagnosed in vivo with 100% accuracy. This was a follow-up study on our previous work focussed on the slowed-down dominant EEG rhythm in AD as compared to FTLD (Goossens et al., 2017). Here, we found that both marker types are able to reveal unique disease properties, which was shown by an improved classification accuracy after combination compared to the accuracy of the separate marker types.

First, we confirmed the value of the established neurochemical biomarkers. Using Aβ_1-42_, T-tau, P-tau_181_ and Nf-L for classification, we achieved a differential AD versus FTLD diagnostic accuracy of 82% in the entire group and nearly 92% in the definite patients. In addition, we found that using the frequency components of the dominant rhythm detected through EEG allowed a classification of 72% in the whole group and 86% in the definite group, which is somewhat behind the neurochemical markers. These accuracies show that both neurochemical markers and EEG markers have a value in distinguishing AD from FTLD and this is a further improvement upon what we found in our previous study on EEG markers only, of which the results were already promising (Goossens et al., 2017). The overall higher classification accuracy on the definite subgroups suggests that clinical misdiagnosis of AD versus FTLD still occurs, despite clinical follow-up that contributed to clinical diagnostic certainty. Therefore, it remains important to investigate additional diagnostic markers that can improve the diagnostic accuracy. The results found in this study are very promising from that perspective.

When we combined the neurochemical and the EEG features, the diagnostic accuracy increased significantly. In the whole group, the accuracy went up from at best 82% (NCM) to almost 85% (p < 10^−15^), while in the definite group only, it went up from at best nearly 92% (NCM) to almost 94% (p = 8.6 x 10^−5^). This suggests the NCM and EEG markers are complementary, revealing different aspects of the disease and therefore confirms again their relevance in developing additional diagnosis tools.

In a number of participants, we detected two dominant EEG peaks. We hypothesise that the mechanism behind the slowing down of the dominant rhythm is not a literal slowing down, but rather a fading out of the normal ground rhythm and the appearance of a new, slower rhythm that starts to dominate the first rhythm. The fact that we only noticed this phenomenon in some of the participants may mean that only they were in a transient state between normal and slowed dominant rhythm. If such a transient state exists, there are only two distinct dominant frequencies in the course of the disease, with a clear transition from the one to the other, i.e. when the slower frequency overtakes the original frequency.

Based on these results, we cannot determine whether everyone goes through this double-peak transition and at what moment in the course of the disease. This would require a longitudinal study, recruiting people who have been diagnosed with AD or FTLD, but who still have a healthy dominant rhythm. If this transition is the true mechanism behind the slowing down of the EEG ground rhythm in AD or FTLD, it probably happens during at a specific disease stage.

In our population, we noticed a few people with no distinguishable dominant peak in many of the channels, most of whom had definite AD. The majority of people had one clear dominant peak in every region and several people had two peaks. In that last group, the patients with AD and two peaks almost all had a probable diagnosis. In addition, there were more FTLD patients with two peaks than AD, especially in the definite group. Based on our data, this suggests that this phenomenon occurs more often in FTLD, which warrants further research.

A possible explanation for having no dominant EEG rhythm could be that the disease has progressed even further such that even the slow rhythm has started to fade out. This is supported by the fact that almost all the AD patients who lacked a clear peak had a definite diagnosis, indicating that disease had progressed to the severe stage. This seemed only valid in the AD group though, as there were no FTLD patients with a definite, and only two with a probable diagnosis who lacked a clear peak in their spectrum. Together with the higher prevalence of a double peak in FTLD, these are interesting new pointers towards more detailed spectral EEG differences between AD and FTLD.

To conclude, we have shown that both neurochemical and EEG biomarkers have very promising value towards differential diagnosis of AD versus FTLD. Combining both biomarker categories resulted in classification improvement as compared to their own diagnostic accuracies. Furthermore, we found evidence that the slowing down of the dominant EEG rhythm might be a gradual appearance of a slow rhythm and fading out of the healthy ground rhythm, rather than a gradual slowing down of the healthy rhythm. Finally, we have discovered two differences in the occurrence of the dominant frequency: people lacking a clear peak almost all had definite AD, while people with two peaks more often had FTLD. These were unexpected findings that are very interesting to investigate further.

## Data Availability

The authors do not have permission to share the data.

## 5. Acknowledgements

This work was funded in part by the University of Antwerp Research Fund and the Institute Born-Bunge (IBB, www.bornbunge.be). INNOTEST hTau-Ag and INNOTEST Phospho-Tau(181P) ELISA kits were kindly provided by Fujirebio Europe, Belgium and Nf-L ELISA kits were kindly discounted by IBL International GmbH, Germany.

